# Association of neighborhood social determinants of health, race and ethnicity, and severe maternal morbidity on the frequency of emergency department visits and preventable emergency department visits among pregnant individuals

**DOI:** 10.1101/2023.08.07.23293765

**Authors:** Richard Holtzclaw, Seuli Bose-Brill, Naleef Fareed

## Abstract

**Background:** The relationship between emergency department (ED) use and SDoH (both individual or neighborhood) factors is complex, and critical factors such as racioethnicity and obstetric comorbidities may moderate this relationship among pregnant individuals. The public health implications of this complex relationship are important for pregnant women because frequent ED visits (both non-preventable and preventable) increase the likelihood of adverse maternal and infant health outcomes and resource burden to communities.

**Methods:** Our quantitative study analyzed clinical, billing, and census data about pregnant individuals from a Tertiary Medical Center (TMC) between 2017 and 2020. To classify visits as preventable, we used an updated New York University ED algorithm. The address of the patient during their ED visit was linked to an area-level deprivation measure to represent neighborhood SDoH. Race and ethnicity data were extracted from the electronic health record and clinical diagnosis data was extracted for obstetric comorbidity ICD-10 codes related to increased risk of severe maternal morbidity (SMM). The date of a clinical diagnosis was used to determine if a specific set of comorbidities were present during a pregnancy. Other sociodemographic and clinical variables were extracted for model adjustment. A negative binomial regression was used to fit the data (n=13,357) to examine the frequency of ED and preventable ED visits based on neighborhood SDoH, race and ethnicity, obstetric comorbidity, and the interactions of these variables.

**Results:** Adjusted model estimates indicated that individuals who identified as non-Hispanic Black experienced higher frequency of ED use across all levels of neighborhood deprivation, and the ED use among non-Hispanic Black individuals in least deprived neighborhoods were higher than or similar to individuals who identified with other racial and ethnic groups who lived in the most deprived neighborhoods. Non-Hispanic Black individuals had the highest frequencies of ED use compared to individuals who identified with other race and ethnic groups whether SMM was present or not, and the frequencies of ED use among non-Hispanic Black individuals with an absence of obstetric comorbidity was higher than individuals who identified with other race and ethnic groups with a comorbidity. Model estimates also indicate that the probabilities of preventable ED visit did not vary race and ethnicity intersected by SDoH. Individuals with obstetric comorbidities had higher probability of a preventable ED visit compared to those not at risk of SMM regardless of different levels of SDoH opportunity. Our study quantifies these differences in estimates between neighborhood SDoH, race and ethnicity, and SMM risk.

**Conclusions:** Examination of interventions to address higher ED use among pregnant individuals require an intersectional lens through which policymakers can gain a nuanced perspective on how ED use is influenced by SDoH, race and ethnicity, and risk of SMM among vulnerable individuals.

## 1.1 Introduction

Prior research has demonstrated that communities with lower neighborhood social determinants of health (SDoH) had higher rates of ED visits (6). There is some evidence to indicate that racioethnic background affects Emergency Department (ED) use (7). These differences are social, not biological. There is, however, a dearth of evidence about the factors that contribute to high ED use among pregnant individuals. If individuals from areas with less opportunity use the ED more among the general population (3,4), similar trends may exist among pregnant individuals. Social determinants of health can be defined as conditions in which people are born, grow, live, work, and age (5). ED use during pregnancy has been tied to adverse outcomes such as preterm delivery (1,2).

The effects of neighborhood SDoH and how they intersect with race and ethnic identity, a measure of individual SDoH is a nuanced assessment of the relationship between SDoH and ED use that has yet to be explored in detail. Individually, research indicates that Non-Hispanic Black individuals are twice as likely to have a prenatal ED visit (8–10), and obstetric comorbidities that are associated with severe maternal morbidity (SMM) conditions have been linked with pre-delivery ED visits (11,12).

The discussion of ED use can also be examined through preventable ED use. Individuals who receive care regularly at the ED often do not have regular doctors or continuity in their care (13). Continuity of care is a vital part of pregnancy increasing quality of care and patient perceptions (14). SDoH, Race and ethnicity, and SMM are interrelated and can influence ED visits among pregnant individuals differentially when considered together. For example, it has been shown that neighborhood SDoH are related to the increased instance of SMM conditions (15). SMM has been found to be more common among racial and ethnic groups other than non-Hispanic White (16). Such associations between race and ethnicity, neighborhood SDoH, and SMM suggest that an intersectional view (17–19) provides an opportunity to examine the complex relationship between ED visits and SDoH among pregnant individuals by providing a nuanced view by which race and ethnicity and SMM intersect with neighborhood SDoH with regard to ED visits.

ED use, and preventable ED use, among pregnant individuals is a critical public health issue because it could be linked to adverse health outcomes among the most vulnerable individuals in society (20), including pregnant individuals from specific racial and ethnic groups, high-deprivation neighborhoods, and those living with chronic conditions. It can also be linked to the use of resources for nonessential treatments or tests, increasing the burden on hospital staff, excessive medical errors, and increased financial burden (1,20). The goal of our study was to investigate the association between two measures of ED use: ED visit frequency and preventable ED use, and the intersection of neighborhood SDoH with race and ethnicity and obstetric comorbidities related to SMM among pregnant individuals at a selected tertiary care center (TCC).

## 2.1 Methods

### 2.1.1 Study Design, setting, and sample

An analytical data set was created using diagnostic and demographic data extracted from the electronic health records (EHR) of our TCC located in the Mid-West United States that provides services across the continuum of care at six hospitals, including general tertiary care and specialty hospitals (i.e., brain and spine, cardiac, and rehabilitation), and multiple specialties including women’s health. Typically, the Obstetrics and Gynecology (OB/GYN) department at the TCC provides prenatal care to over 5,000 individuals, including those with high-risk pregnancies. Inclusion Criteria were individual pregnancy episodes of pregnant individuals who were at least 18 years of age at time of delivery and delivered at the TCC between 2017 and 2020. An episode refers to a unique pregnancy for which prenatal care was received at the TCC. We only include pregnancies that came to term to ensure comparison of ED use over the full course of pregnancy, and not introduce observations with ectopic pregnancies, spontaneous abortions, or other conditions that may have a complex relationship with ED use. Deliveries outside the TCC were not included in the analysis.

## 3.1 Ethics

Retrospective data were obtained for individual encounters from the TCC’s Information Warehouse (IW) in accordance with the Honest Broker protocol and our study was approved by the Biomedical IRB at [Blinded]

## 4.1 Data

The analytical data set was created using multiple data sources: clinical discharge data from the EHR, billing data from revenue cycle, and neighborhood SDoH data using the [Blinded]. Data were aggregated to the pregnancy episode level.

Separate files of EHR data were provided by the TCC Information Warehouse for demographic data, delivery date, address during delivery, gestational length, discharge data, neighborhood SDoH, and diagnoses. These files were combined into one analytical file by matching a unique medical record number (MRN), pregnancy episode, and ZIP Code. First the address during delivery, delivery date, and gestational data files were combined using MRN and delivery dates. These dates were used to determine pregnancy episode. Patient demographics were then matched to each episode by MRN to form a patient profile data set. Pregnancy episode was calculated in the diagnostic data file by date range, then this data was merged with the patient profile by MRN and episode to create a diagnostic profile. Separately, the discharge data file also had the pregnancy episode determined by date range and was combined with the patient profile by MRN and episode number. This new file was used to calculate ED visit and preventable ED frequencies, by episode and ZIP Code. That frequency along with insurance information was then merged with the diagnostic profile by MRN and episode number. Finally, the [Blinded] was merged by the ZIP Code of the patient during the pregnancy episode. This provided a single analytical file with all the information needed for this evaluation.

### 4.1.1 Exposure, outcome, and covariates

The primary exposures were neighborhood-level SDoH, race and ethnicity, and presence of obstetric comorbidities. Neighborhood SDoH was operationalized using the [Blinded] ([BLINDED]). The [BLINDED] is a geographically based measure involving the following area level SDoH factors: income, employment, transportation, crime, health, education, and housing quality. The residential ZIP Code of a patient at delivery was assigned an [BLINDED] score. This index was developed by researchers at the TCC to standardize a measure of neighborhood opportunity of development and well-being across the state. The [BLINDED] scores were converted to tertiles based on the state distribution of [BLINDED] scores for this analysis. Higher tertiles represent higher opportunity for individuals in the corresponding ZIP Codes. More information about the development and use of [BLINDED] can be found in the study by Fareed and colleagues (21). Race and ethnicity is operationalized as non-Hispanic White, Non-Hispanic Black, and Hispanic. Finally, a binary representation of presence of obstetric comorbidities was constructed. SMM risk was considered present if any of the underlying comorbidities from the obstetric comorbidities outlined by Leonard and colleagues (22) was present and absent otherwise. These obstetric comorbidities include preexisting conditions such as diabetes, asthma, and hypertension, as well as pregnancy-related conditions such as placenta accreta, gestational diabetes mellitus, and placenta previa (please see Appendix 1 for a list of International Classification of Diseases, Tenth Revision [ICD-10] codes for the clinical comorbidities used in our study). Leonard and colleagues’ research shows the presence of these obstetric comorbidities increase the risk of SMM in pregnant individuals. In taking an intersectional view, we examined the two-way interactions of these three exposures.

The primary outcomes of interest were the frequency of ED visits by an individual during a pregnancy episode and preventable ED visits as defined using the NYU ED algorithm. The ICD-10 codes were used to identify diagnoses and to characterize ED visits by specific clinical diagnoses and find ED visits that were preventable. To classify visits as preventable, we used the updated New York University (NYU) ED algorithm (13,23). This previously validated algorithm assigns probabilities and classifies each ED visit as urgent (emergent-not preventable/avoidable: immediate care in an ED setting needed and the condition could not have been prevented/avoided with ambulatory care, such as heavy or ongoing bleeding, passing tissue from the vagina, abdominal pain, protracted nausea or vomiting), emergent but preventable or avoidable (immediate care in an ED setting needed but the condition could have been prevented or avoided with timely and effective ambulatory care, such as dehydration, asthma with acute exacerbation, diabetes with hyper – or hypoglycemia), emergent but primary care treatable (care is needed within 12 hours but could be provided in a primary care setting, such as epigastric pain, headache; including migraine), and non-emergent (immediate care not required within 12 hours, such as headache, cough, low back pain, fatigue and weakness). We created a single preventable ED variable that was coded as one if the probability of an ED event from any of the four categories was higher than 50% and zero otherwise, as suggested by prior literature (24).

Individual socio-demographic and clinical factors may confound our estimated coefficients as demonstrated in prior literature. For example, individuals who are enrolled in Medicaid may have more ED visits and more individuals enrolled in Medicaid might live in more deprived neighborhoods, identify as non-Hispanic Black, or have one or more obstetric comorbidities. Maternal age at delivery was used because it can be associated with additional complications associated with ED use (25) and certain facets of neighborhood SDoH impact younger age groups more (26). Maternal age was categorized as 18-24; 25-34; 35 and above. Individuals in areas of neighborhood SDoH related to less opportunity have a higher chance of being on Medicaid, and those who are insured by Medicaid seek care in the ED more frequently (26–28). Insurance status was included separating Medicaid from private insurance and treating it as a categorical variable (private insurance, Medicaid, other insurance statuses such as Medicare, uninsured, etc). Gravidity allows for controlling for the number of past pregnancies as some studies have suggested gravidity might be associated with increased ED use (2) and individuals who have lower gravidity may live in specific neighborhoods or identify with a specific race and have systematically different obstetric comorbidities present. Pregnancy episode was measured as a count for the number of episodes, which allows for each pregnancy to be accounted for as a unique experience, which may have differing ED use and be influenced by race and ethnicity, neighborhood SDoH, and obstetric comorbidities related to SMM that exist during the pregnancy. We also use binary year variables (2017 reference) to account for historical trend effects with ED visits.

## 5.1 Statistical analysis

A negative binomial regression model was used to obtain estimated incidence rate ratios (IRR) for ED visit frequency. In addition, a logistic regression was used to find the Odds Ratios (OR) for preventable ED visits. The first specifications tested the relationship between SDoH and the primary outcomes and the second specification tested interactions between neighborhood SDoH and race and ethnicity and neighborhood SDoH and presence of obstetric comorbidities and the association with the primary outcomes. Covariates were included based on a directly acyclic graph: maternal age at delivery, insurance status, parity, pregnancy episode number, and study year. For the negative binomial regression random effects was used to adjust estimates and provide robust standard errors based on multiple pregnancies per individual. An offset was incorporated to account for varying gestational lengths. Adjusted predicted means were obtained from both model specifications. All analyses were conducted using Stata, version 17.0 (StataCorp).

## 6.1 Results

Participant characteristics are provided in Table 1. Complete case wise deletion was used in the analytical data set. Overall, 50 or 0.3% of observations were dropped. Of those, 49 had no date of birth and maternal age could not be calculated, and one observation had no recorded ZIP Code. Table 1 describes the demographics of the study sample. The total sample size was 14,340 pregnancy episodes with about a third living in locations with poor neighborhood SDoH (i.e., Tertile 1), 87% of whom sought care for at least one pregnancy episode, and 61% who identified as non-Hispanic White. More than 58% had the presence of one or more obstetric comorbidities, about 37% were enrolled on Medicaid, and the mean maternal age was 30 at delivery.

**Table 1.**
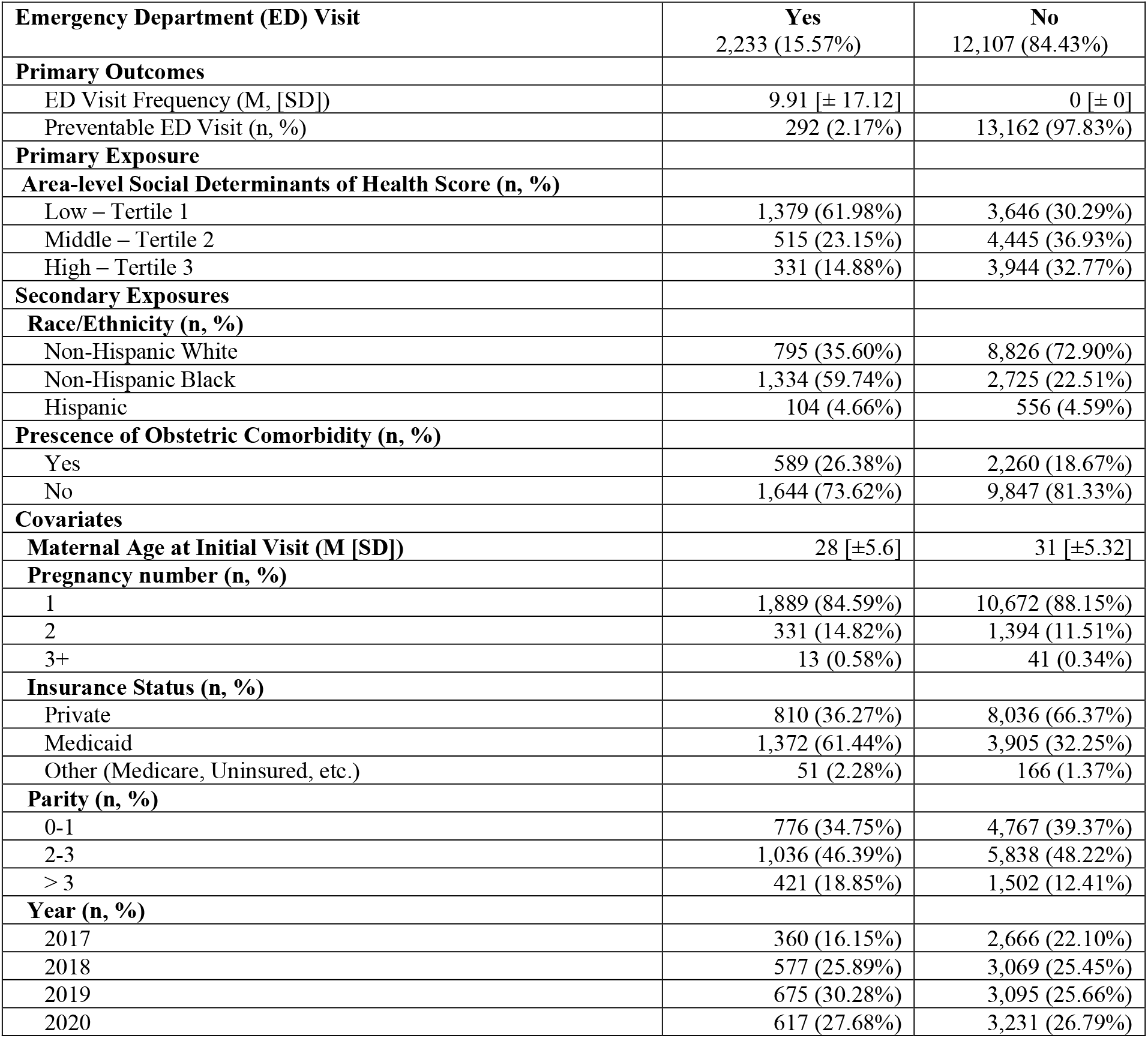
Characteristics of individual pregnancy episodes included in the study sample (N=13,357)

Table 2 shows the estimated IRRs for the frequency of ED visits and the estimated ORs for preventable ED visits among pregnant individuals. The estimated differences in ED visit frequency differed by the primary variable adjusted for our covariates. Race and ethnicity had significant effects on frequency of ED visits. Non-Hispanic Black individuals utilized the ED during pregnancy almost 9 times as much as non-Hispanic White individuals. Hispanic individuals frequented the ED almost three times as much as their White counterparts. Regarding neighborhood SDoH, those from more vulnerable areas frequented the ED more often. Individuals from neighborhoods with the most opportunity, as calculated by the [BLINDED], generally had approximately two fifths the ED visit frequency as those in neighborhoods with the least opportunity. For preventable ED visits, presence of obstetric comorbidities had significant effects on preventable ED visits. The odds of a pregnant individual with obstetric comorbidities visiting the ED with one of the diagnoses categorized as preventable is over 18 times higher than the odds of a preventable ED visit without obstetric comorbidities.

**Table 2.** Adjusted Incident Rate Ratios for frequency of ED visit and preventable ED visits with neighborhood SDoH complete dataset (2017-2020)

|  | **ED Visit Frequency** |  | **Preventable ED Visits** |  |
| --- | --- | --- | --- | --- |
| **Variable** | **Unadj.IRR (95% CI)**<br>**(n = 14,340)** | **Adj.IRR (95% CI)**<br>**(n = 14,340)** | **Unadj.OR (95% CI)**<br>**(n = 14,340)** | **Adj.OR (95% CI)**<br>**(n = 14,340)** |
| **SDoH (Area level measure)** |  |  |  |  |
| Tertile 1 (lowest opportunity) | Reference | Reference | Reference | Reference |
| Tertile 2 | 0.15*** (0.12 - 0.18) | 0.49*** (0.35 - 0.7) | 0.71** (0.65 - 1.19) | 0.87 (0.65 - 1.16) |
| Tertile 3 (highest opportunity) | 0.08*** (0.06 - 0.1) | 0.42*** (0.29 - 0.64) | 0.49*** (0.39 - 0.85) | 0.85 (0.58 - 1.23) |
| **Race and Ethnicity** |  |  |  |  |
| Non-Hispanic White | Reference | Reference | Reference | Reference |
| Non-Hispanic Black | N/A | 9.08*** (7.0 - 11.77) | N/A | 1.09 (0.81 - 1.46) |
| Hispanic | N/A | 2.71*** (1.62 - 4.55) | N/A | 0.72 (0.38 - 1.28) |
| **Obstetric Comorbidity** |  |  |  |  |
| Absent | Reference | Reference | Reference | Reference |
| Present | N/A | 1.5*** (1.19 - 1.9) | N/A | 19.49*** (12.73 - 29.83) |

**Footnote:** \*\*\*p<0.01 Adj.IRR for maternal age, insurance status, parity, general ob visits, delivery year.

Based on the predicted means from the adjusted interaction model, non-Hispanic Black individuals in the most deprived neighborhoods had higher mean ED visit frequency among all individuals. There were no statistically significant differences across racial/ethnic groups for preventable ED visit frequency (**Figure 1**).

**Figure 1.**
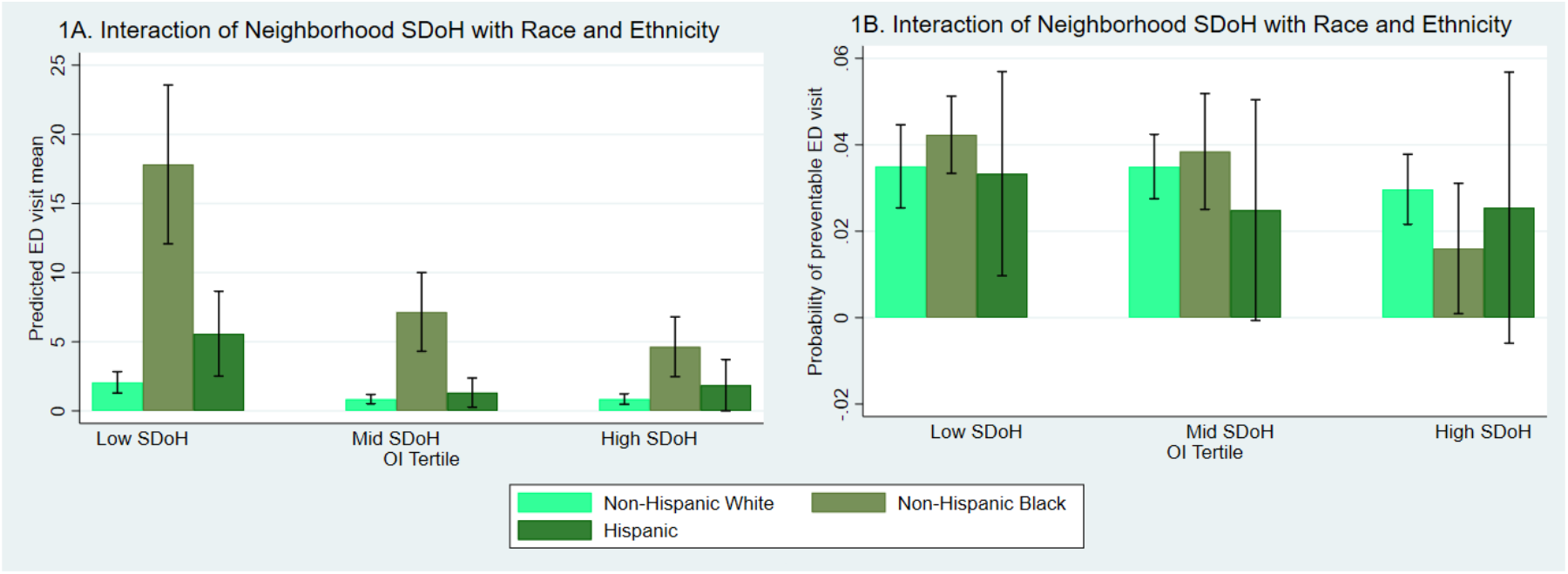
**(A&B)**. Interaction plots of social determinants of health (SDoH) and race and ethnicity for **A**) predicted Emergency Department (ED) visit frequency and **B)** probability of preventable ED visit.

Individuals with obstetric comorbidities had higher mean ED visit frequency compared to those not at risk of SMM across neighborhoods of different levels of SDoH opportunity. Individuals with obstetric comorbidities had significantly higher probabilities of a preventable ED visit compared to those not at risk of SMM regardless of different levels of SDoH opportunity (**Figure 2**).

**Figure 2.**
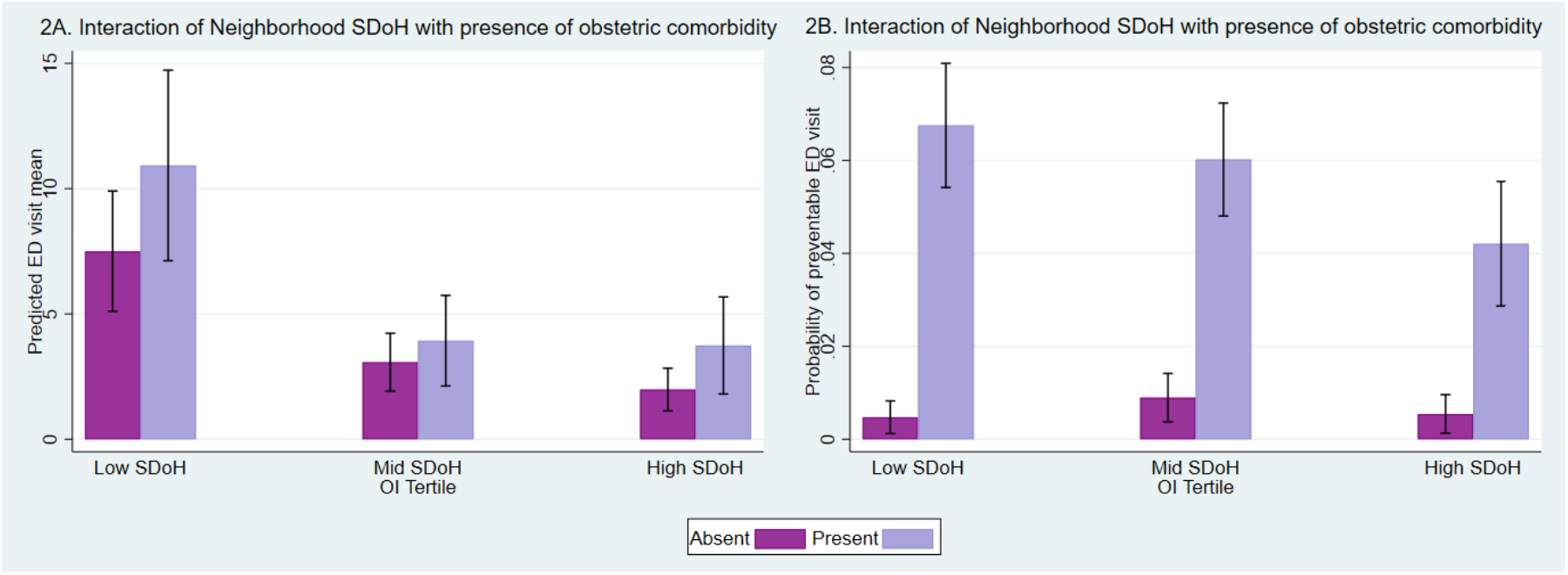
**(A & B)** Interaction plots of social determinants of health (SDoH) and presence of obstetric comorbidity for **A**) predicted Emergency Department (ED) visit frequency and **B)** probability of preventable ED visit.

## 7.1 Discussion

Previous literature indicated that neighborhood SDoH greatly impacts the choice to visit the ED. Proximity to a care facility or ability to travel are some of the biggest factors (15). The results of our study might indicate these same factors influence pregnant individuals use of the ED. For example, we found many of the neighborhoods with high ED visit frequency lacked access to a vehicle. People who reside in highly deprived neighborhoods are more likely to visit the ED than those who live in wealthy areas, regardless of race and ethnic background or presence of one or more obstetric comorbidities related to SMM. Also, our results disconcertingly indicate that pregnant individuals who identify as non-Hispanic Black utilized the ED significantly more than even those from the most deprived areas.

Our study indicates that pregnant individuals align with the general population in which previous research demonstrates that individuals on Medicaid use the ED more frequently (29,30). A national study looking at ED use by individuals with commercial insurance found that 17% of pregnant women visited the ED at least once (31). Our study’s population was a little over one percentage point lower, at 15.96%.

The areas we examined of the highest ED frequency and lowest neighborhood SDoH represented neighborhoods designated as high priority in regards to infant and maternal health (32). Our study found that these areas currently see high rates of preventable ED use. If pregnant individuals in primarily non-Hispanic Black neighborhoods do not receive primary care due to distance, resource barriers, or discrimination this may result in increased ED visit frequency and maternal near misses. A maternal near miss is defined as “a woman who nearly died but survived a complication that occurred during pregnancy, childbirth, or within 42 days of termination of pregnancy” (33). These near misses are especially common among non-Hispanic Black individuals, and often result when ongoing conditions escalate to the case of an emergency (33).

While the effect of neighborhood deprivation on preventable ED use is less extreme, the presence of obstetric comorbidities seem to have a great effect on preventable ED visits with those at risk of SMM having much higher odds of preventable ED visits. High rates of ED use for primary care can result in an increase of preventable ED visits and a reduction in care continuity and quality of care during pregnancy increasing the high risk individuals with obstetric comorbidities already have of maternal complication including death (12).

A 2022 study suggests more robust measures of SDoH in the EHR often have missing values that influence the outcome of predictive models (34). Our study uses SDoH measures external to the EHR, looking at the neighborhood variation where patients reside. This should help mitigate the effect of missing data and possibly demonstrate the value of more robust data collection to improve the effectiveness of the EHR in predicting ED frequency or preventable visits.

## 8.1 Limitations

Only data from the selected TCC were obtained. We were not able to capture data outside of our TCC, a data gap that has implications for our results. The assumption was made that during a single pregnancy, care would be sought within the same network.

Extraction directly from the EHR data warehouse to measure conditions is not as good as chart abstraction. There are concerns of data variability surrounding obstetrics data from EHRs (35). However, EHR data allows for ease of collection of multiple types of information including demographic, diagnostic, and clinical data. It also allows for the connection of non-pregnancy related visits and diagnoses during pregnancy. EHR data is also easily deidentified to protect the identity of those included in our study.

SMM measurement is complicated, and our estimates are conservative. Presence of obstetric comorbidity related to SMM was assumed if a diagnosis of past or current obstetric comorbidities were recorded during or before pregnancy. These risk factors associated with SMM may not have been reported for each episode in which they exist or may have been captured at other facilities and not recorded in the TCC’s EHR data warehouse.

Separating out the neighborhood SDoH allows for examining how much of an effect the complex social and physical environments have on individuals, which may not be captured by individual SDoH factors. Our method excludes individual SDoH factors, and little to no previous literature exists on the relationship between ED use and these individual factors. It is possible these factors have an effect either on their own or related to neighborhood SDoH.

## 9.1 Conclusion

Pregnant individuals that use the ED more frequently are already part of vulnerable populations, being individuals from minoritized groups, in areas of lower opportunity and fewer resources. In addition, those with one or more obstetric comorbidities present during their pregnancy have increased instances of preventable ED use. Although race and ethnicity was the most influential predictor of ED use, the underlying relationships within neighborhood SDoH, race and ethnicity, and obstetric comorbidity result in even more frequent ED use and preventable ED visits, while controlling for other possible factors. Examination of interventions to address higher ED use and avoid preventable ED visits among pregnant individuals requires an intersectional lens through which policymakers can gain a nuanced perspective on how ED use is influenced by SDoH, race and ethnicity, and obstetric comorbidity among vulnerable individuals.

## Data Availability

Data used in the study analysis is subject to restrictions as determined by the institutional IRB.

## Author Declarations

### Footnote

Social Determinants of Health (SDoH); Emergency Department (ED); Severe Maternal Morbidity (SMM); Tertiary Care Center (TCC); Electronic Health Records (EHR); Medical Record Number (MRN); International Classification of Diseases, Tenth Revision (ICD-10); New York University (NYU)

I confirm all relevant ethical guidelines have been followed, and any necessary IRB and/or ethics committee approvals have been obtained.

Yes

The details of the IRB/oversight body that provided approval or exemption for the research described are given below:

Retrospective data were obtained for patient encounters from our AMC’s Information Warehouse (IW) in accordance with the Honest Broker protocol. The study protocol was approved by the Ohio State University Institutional Review Board (IRB).

All necessary patient/participant consent has been obtained and the appropriate institutional forms have been archived.

Yes

I understand that all clinical trials and any other prospective interventional studies must be registered with an ICMJE-approved registry, such as ClinicalTrials.gov. I confirm that any such study reported in the manuscript has been registered and the trial registration ID is provided (note: if posting a prospective study registered retrospectively, please provide a statement in the trial ID field explaining why the study was not registered in advance).

Study uses retrospective data.

I have followed all appropriate research reporting guidelines, such as any relevant EQUATOR Network research reporting checklist(s) and other pertinent material, if applicable.

Yes

### Appendix

#### Appendix 1

##### ICD-10 codes used in Analyses

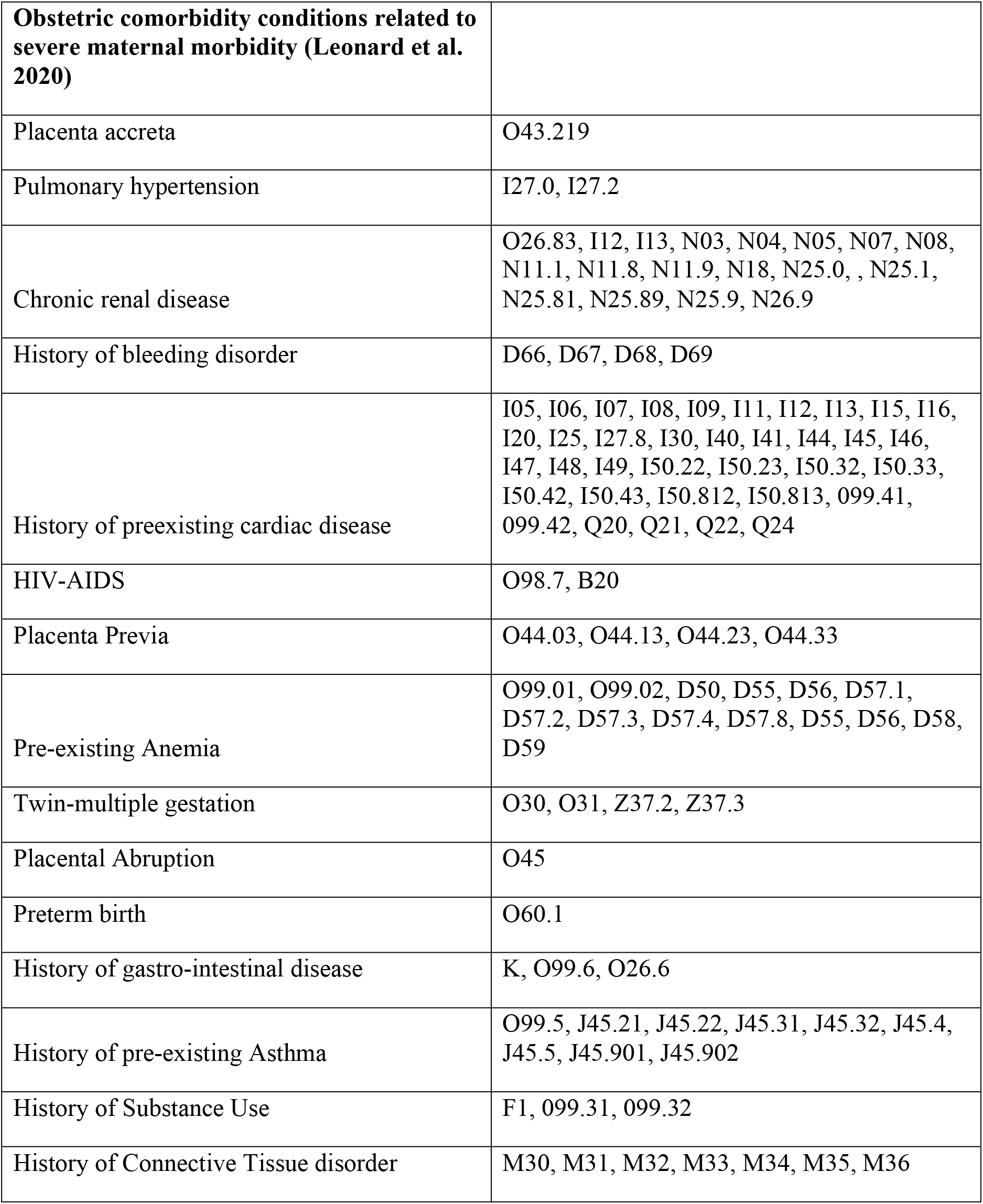
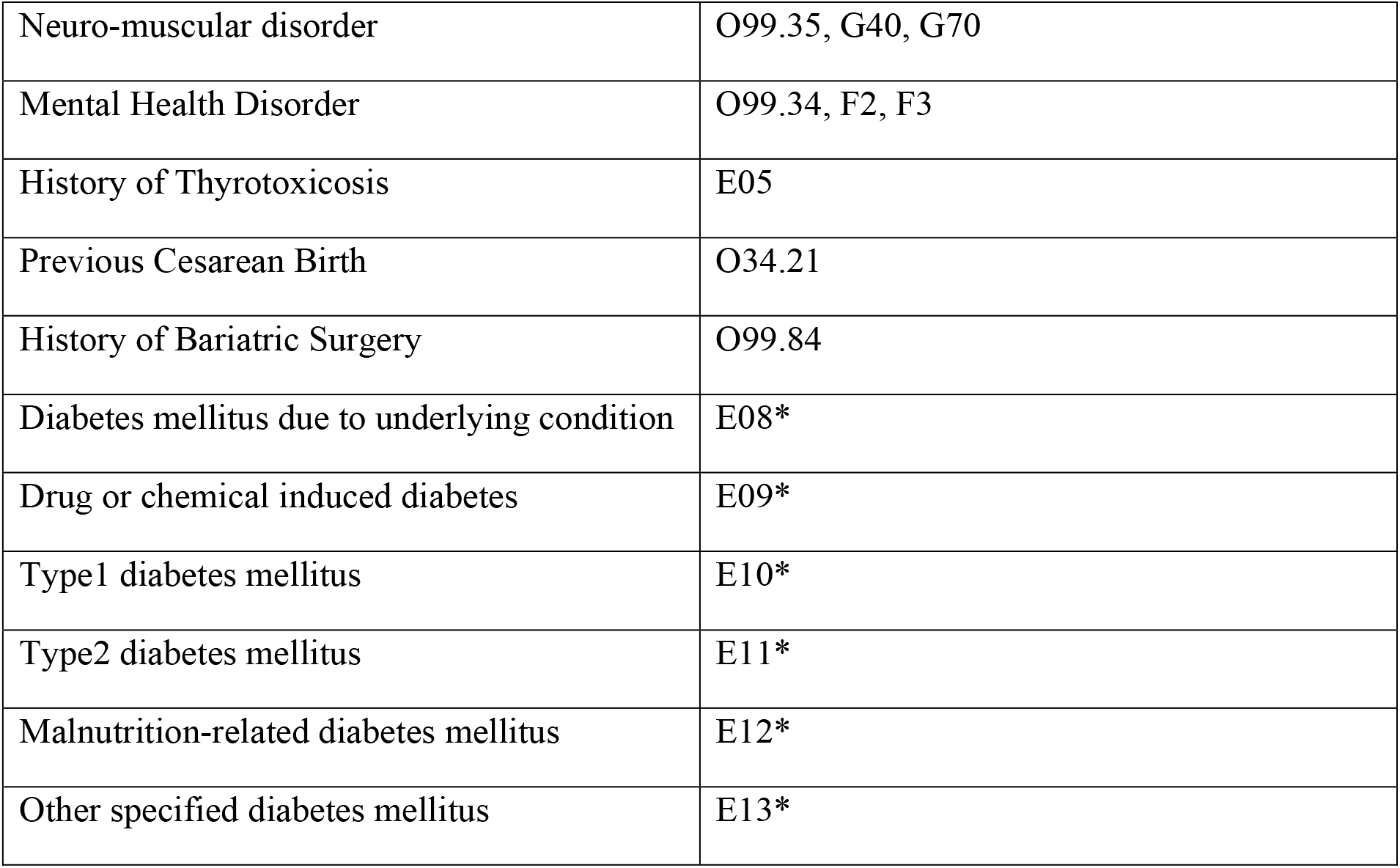

## Bibliography

1. Magriples U, Kershaw TS, Rising SS, Massey Z, Ickovics JR. Prenatal health care beyond the obstetrics service: Utilization and predictors of unscheduled care. Am J Obstet Gynecol. 2008 Jan;198(1):75.e1-7.

2. Varner CE, Park AL, Ray JG. Prepregnancy emergency department use and risks of severe maternal and neonatal morbidity in canada. JAMA Netw Open. 2022 Sep 1;5(9):e2229532.

3. Doran KM, Kunzler NM, Mijanovich T, Lang SW, Rubin A, Testa PA, et al. Homelessness and other social determinants of health among emergency department patients. J Soc Distress Homeless. 2016 Jul 2;25(2):71–7.

4. Singh GK, Daus GP, Allender M, Ramey CT, Martin EK, Perry C, et al. Social Determinants of Health in the United States: Addressing Major Health Inequality Trends for the Nation, 1935-2016. Int J MCH AIDS. 2017;6(2):139–64.

5. W H O. About social determinants of health [Internet]. [cited 2020 Apr 15]. Available from: https://www.who.int/social_determinants/sdh_definition/en/

6. McIntire RK, Romney MC, Alonzo G, Hutt J, Bartolome L, Wood G, et al. Do Employees From Less-Healthy Communities Use More Care and Cost More? Seeking to Establish a Business Case for Investment in Community Health. Prev Chronic Dis. 2019 Jul 25;16:E95.

7. Parast L, Mathews M, Martino S, Lehrman WG, Stark D, Elliott MN. Racial/ethnic differences in emergency department utilization and experience. J Gen Intern Med. 2022 Jan;37(1):49–56.

8. Harrell T, Howell EA, Balbierz A, Guel L, Pena J, Janevic T, et al. Improving Postpartum Care: Identifying Opportunities to Reduce Postpartum Emergency Room Visits Among Publicly-Insured Women of Color. Matern Child Health J. 2022 Apr;26(4):913–22.

9. Enriquez R, Griffin MR, Carroll KN, Wu P, Cooper WO, Gebretsadik T, et al. Effect of maternal asthma and asthma control on pregnancy and perinatal outcomes. J Allergy Clin Immunol. 2007 Sep;120(3):625–30.

10. Brown E Richard, Ojeda V, Wyn R, Levan R. Racial and Ethnic Disparities in Access to Health Insurance and Health Care [Internet]. UCLA Center for Health Policy Research; 2000 Apr [cited 2022 Aug 10]. Available from: https://www.kff.org/wp-content/uploads/2013/01/racial-and-ethnic-disparities-in-access-to-health-insurance-and-health-care-report.pdf

11. Lazariu V, Nguyen T, McNutt L-A, Jeffrey J, Kacica M. Severe maternal morbidity: A population-based study of an expanded measure and associated factors. PLoS ONE. 2017 Aug 7;12(8):e0182343.

12. Alford JM, Williams SN, Oriaku MN, White D, Schwartzman A, Jackson G. National hospital care survey demonstration projects: severe maternal morbidity in inpatient and emergency departments. Natl Health Stat Report. 2021 Oct;(166):1–15.

13. Billings J, Parikh N, Mijanovich T. Emergency department use in New York City: a substitute for primary care? Issue Brief (Commonw Fund). 2000 Nov;(433):1–5.

14. Perdok H, Verhoeven CJ, van Dillen J, Schuitmaker TJ, Hoogendoorn K, Colli J, et al. Continuity of care is an important and distinct aspect of childbirth experience: findings of a survey evaluating experienced continuity of care, experienced quality of care and women’s perception of labor. BMC Pregnancy Childbirth. 2018 Jan 8;18(1):13.

15. [Blinded for review]

16. Mujahid MS, Kan P, Leonard SA, Hailu EM, Wall-Wieler E, Abrams B, et al. Birth hospital and racial and ethnic differences in severe maternal morbidity in the state of California. Am J Obstet Gynecol. 2021 Feb;224(2):219.e1–219.e15.

17. Bower KM, Thorpe RJ, Rohde C, Gaskin DJ. The intersection of neighborhood racial segregation, poverty, and urbanicity and its impact on food store availability in the United States. Prev Med. 2014 Jan;58:33–9.

18. Hung P, Liu J, Norregaard C, Shih Y, Liang C, Zhang J, et al. Analysis of Residential Segregation and Racial and Ethnic Disparities in Severe Maternal Morbidity Before and During the COVID-19 Pandemic. JAMA Netw Open. 2022 Oct 3;5(10):e2237711.

19. Interrante JD, Tuttle MS, Admon LK, Kozhimannil KB. Severe maternal morbidity and mortality risk at the intersection of rurality, race and ethnicity, and medicaid. Women’s Health Issues. 2022 Jun 25;32(6):540–9.

20. Trzeciak S, Rivers EP. Emergency department overcrowding in the United States: an emerging threat to patient safety and public health. Emerg Med J. 2003 Sep;20(5):402–5.

21. Fareed N, Swoboda CM, Jonnalagadda P, Griesenbrock T, Gureddygari HR, Aldrich A. Visualizing Opportunity Index Data Using a Dashboard Application: A Tool to Communicate Infant Mortality-Based Area Deprivation Index Information. Appl Clin Inform. 2020 Aug 5;11(4):515–27.

22. Leonard SA, Kennedy CJ, Carmichael SL, Lyell DJ, Main EK. An expanded obstetric comorbidity scoring system for predicting severe maternal morbidity. Obstet Gynecol. 2020 Sep;136(3):440–9.

23. Johnston KJ, Allen L, Melanson TA, Pitts SR. A “patch” to the NYU emergency department visit algorithm. Health Serv Res. 2017 Aug;52(4):1264–76.

24. Ballard DW, Price M, Fung V, Brand R, Reed ME, Fireman B, et al. Validation of an algorithm for categorizing the severity of hospital emergency department visits. Med Care. 2010 Jan;48(1):58–63.

25. Cavazos-Rehg PA, Krauss MJ, Spitznagel EL, Bommarito K, Madden T, Olsen MA, et al. Maternal age and risk of labor and delivery complications. Matern Child Health J. 2015 Jun;19(6):1202–11.

26. Dwyer RE, Neilson LA, Nau M, Hodson R. Mortgage worries: young adults and the US housing crisis. Socio-Economic Review. 2016 Jul;14(3):483–505.

27. Kim H, McConnell KJ, Sun BC. Comparing Emergency Department Use Among Medicaid and Commercial Patients Using All-Payer All-Claims Data. Popul Health Manag. 2017 Aug;20(4):271–7.

28. Hunt KA, Weber EJ, Showstack JA, Colby DC, Callaham ML. Characteristics of frequent users of emergency departments. Ann Emerg Med. 2006 Jul;48(1):1–8.

29. Taylor YJ, Liu TL, Howell EA. Insurance Differences in Preventive Care Use and Adverse Birth Outcomes Among Pregnant Women in a Medicaid Nonexpansion State: A Retrospective Cohort Study. J Womens Health (Larchmt). 2020;29(1):29–37.

30. Kilfoyle KA, Vrees R, Raker CA, Matteson KA. Nonurgent and urgent emergency department use during pregnancy: an observational study. Am J Obstet Gynecol. 2017 Feb;216(2):181.e1-181.e7.

31. Cunningham SD, Magriples U, Thomas JL, Kozhimannil KB, Herrera C, Barrette E, et al. Association Between Maternal Comorbidities and Emergency Department Use Among a National Sample of Commercially Insured Pregnant Women. Acad Emerg Med. 2017 Aug;24(8):940–7.

32. [Blinded for review]

33. Say L, Souza JP, Pattinson RC, WHO working group on Maternal Mortality and Morbidity classifications. Maternal near miss--towards a standard tool for monitoring quality of maternal health care. Best Pract Res Clin Obstet Gynaecol. 2009 Jun;23(3):287–96.

34. Hall AG, Davlyatov GK, Orewa GN, Mehta TS, Feldman SS. Multiple Electronic Health Record-Based Measures of Social Determinants of Health to Predict Return to the Emergency Department Following Discharge. Popul Health Manag. 2022 Dec;25(6):771–80.

35. Altman MR, Colorafi K, Daratha KB. The reliability of electronic health record data used for obstetrical research. Appl Clin Inform. 2018 Jan;9(1):156–62.

